# Comparison of kinetics of immune responses to SARS-CoV-2 proteins in individuals with varying severity of infection and following a single dose of the AZD1222

**DOI:** 10.1101/2021.07.14.21260510

**Authors:** Deshni Jayathilaka, Chandima Jeewandara, Laksiri Gomes, Tibutius Thanesh Pramanayagam Jayadas, Achala Kamaladasa, Dinuka Guruge, Pradeep Darshana, Thushali Ranasinghe, Inoka Sepali Abyrathna, Saubhagya Danasekara, Buddini Gunathilaka, Heshan Kuruppu, Ananda Wijewickrama, Ruwan Wijayamuni, Lisa Schimanski, T.K. Tan, Graham S. Ogg, Alain Townsend, Gathsaurie Neelika Malavige

**Author notes:** Correspondence should be addressed to: Prof. Neelika Malavige DPhil (Oxon), FRCP (Lond), FRCPath (UK), Allergy Immunology and Cell Biology Unit, Department of Immunology and Molecular Medicine, Faculty of Medical Sciences, University of Sri Jayawardanapura, Sri Lanka. Contributed equally (Deshni Jayathilaka, Chandima Jeewandara).

## Abstract

**Background:** While there have been many studies characterizing the IgG and IgA responses to different SARS-CoV-2 proteins in individuals with natural infection, the induction of IgG and IgA to different viral proteins in vaccinees have not been extensively studied. Therefore, we sought to investigate the antibody responses to SARS-CoV-2 following natural infection and following a single dose of AZD2221, in Sri Lankan individuals.

**Methods:** Using Luminex assays, we characterized the IgG and IgA responses in patients with varying severity of illness and following a single dose of the vaccine at 4 weeks and 12 weeks since onset of illness or following vaccination. Haemagglutination test (HAT) was used to assess the antibodies to the receptor binding domain of SARS-CoV-2 wild type (WT), B.1.1.7, B.1.351 and B.1.617.2 (VOCs) and surrogate neutralizing test to measure ACE2 receptor blocking antibodies.

**Results:** Those with mild illness and in vaccinees, the IgG responses to S1, S2, RBD and N protein increased from 4 weeks to 12 weeks, while it remained unchanged in those with moderate/severe illness. Those who had a febrile illness in 2017 and 2018 (controls) also gave IgG and IgA high responses to the S2 subunit. In the vaccinees, the most significant rise was seen for the IgG antibodies to the S2 subunit (p<0.0001). Vaccinees had several fold lower IgA antibodies to all the SARS-CoV-2 proteins tested than those with mild and moderate/severe illness at 4 weeks and 12 weeks. At 12 weeks the HAT titres were significantly lower to the B.1.1.7 in vaccinees and significantly lower in those with mild illness, and in vaccinees to B.1.351 and for B.1.617.2. No such difference was seen in those with moderate/severe illness.

**Conclusions:** Vaccinees had significantly less IgA to SARS-CoV-2, but comparable IgG responses to those with natural infection. However, following a single dose, vaccinees had reduced antibody levels to the variants of concern (VOC), which further declined with time, compared to natural infection.

## Introduction

The COVID-19 pandemic due to SARS-CoV-2, continues to cause significant mortality and morbidity, and many countries are currently experiencing a worse situation, than at the beginning of the pandemic^1^. Emergence of SARS-CoV-2 variants of concern such as the B.1.1.7 (alpha) and more recently B.1.617.2 (delta) has led to exponential increase of the number of COVID-19 cases and deaths in many countries^1-3^. While the higher income countries have vaccinated a large proportion of their population, resulting in lower hospitalizations and deaths, many lower income and lower-middle income countries are grappling with the increase in the case loads, overburdening of health care resources and the inability to secure adequate doses of COVID-19 vaccines^4^.

Although the duration of protection against re-infection from SARS-CoV-2 in not known, it has been shown that re-infection does occur, especially among older individuals, probably due to waning of immunity^5^. Re-infection has shown to occur particularly with certain variants such as P.1 (gamma) variant in Brazil despite a very high seroprevalence^6^, and also with B.1.351 (beta) due to escape from natural and vaccine induced immunity^7^. Individuals who had experienced milder illness have shown to have reduced levels of neutralizing antibodies compared to those who had severe illness^8,9^. Apart from the presence of neutralizing antibodies to the receptor binding domain (RBD), antibodies specific to S2 and N protein of SARS-CoV-2 are also detected in patients who have recovered from COVID-19^10^. Although the usefulness of antibodies directed against S1, S2 and N protein in preventing re-infection are not known, although IgG and IgA specific to S1, S2 have been detected in breast milk of infected mothers and therefore, possibly provide protection to the neonate^11^. Antibodies against the S2 subunit have been detected in unexposed individuals and S1, S2 and N protein specific memory B cell responses have been detected in those who were infected with SARS-CoV-2^12^. Children and adolescents who were unexposed to SARS-CoV-2 were shown to have a higher frequency of pre-existing IgG antibodies specific to S2, which were able to cross neutralize SARS-CoV-2^13^. The presence of high levels of cross-reactive antibodies to the S2 in children and adolescents have been speculated to reduce disease severity when infected with SARS-CoV-2^13,14^. Although many studies have investigated the role of SARS-CoV-2 specific IgG responses, virus specific IgA was detected during early illness and was shown to be able to neutralize the SARS-CoV-2 virus to a greater extent than virus specific IgG^15^. However, adults with severe illness had higher levels of SARS-CoV-2 specific IgA levels compared to adults with milder illness and children, and was shown to enhance neutrophil activation in vitro and thus release of inflammatory mediators^16^. Therefore, although virus specific IgA is an important component of mucosal immunity, its role in protection vs disease pathogenesis is not clear.

Currently there are several vaccines for COVID-19, which have shown to be safe and have high efficacy rates against the original Wuhan SARS-CoV-2 virus and variable efficacy against variants of concerm^17-19^. However, due to non-availability of adequate quantity of vaccines and also in order to vaccinate as many individuals as fast as possible, some countries have increased the gap between the two doses of vaccine such as AZD2221 to 12 or 16 weeks^20^. While there have been many studies characterizing the IgG and IgA responses to different SARS-CoV-2 proteins in individuals with natural infection, the induction of IgG and IgA to different viral proteins in vaccinees have not been extensively studied. It was recently shown that the mRNA vaccines induce high levels of both IgG and IgA antibodies against the spike protein^21^. However, there are limited data characterizing the IgG, IgA, ACE2-receptor blocking antibodies in individuals with varying severity of natural infection over time, in comparison to those who have received a single dose of the AZD2221 vaccine. Therefore in this study, we investigated the antibody responses in those with varying severity of natural infection and in those who received a single dose of the AZD2221 at 4 weeks and 12 weeks to the S1, S2, RBD and N proteins and also for SARS-CoV-2 variants of concern in a Sri Lankan population.

## Methods

### Patients

Patients confirmed with SARS-CoV2 infection based on the positive RT-PCR who were admitted to a COVID-19 treatment hospital, from the National Institute of Infectious Diseases (NIID), Sri Lanka. They were followed throughout their illness while they were in hospital and the severity grading was based on the worst severity while in hospital. Clinical disease severity was classified as mild, moderate and severe according to the WHO guidance on COVID-19 disease severity ^22^. For this study we recruited two cohorts of patients. Serum samples from the patient cohort 1 (n=30) was used to determine the IgG and IgA antibody levels at 4 weeks since onset of illness, the ACE2 receptor blocking antibody levels and the antibodies to RBD by the HAT assay for the wild type (WT) and SARS-CoV-2 variants. The duration of illness was defined from the day of onset of symptoms and not the day of PCR positivity or admission to hospital. Based on the WHO COVID-19 disease classification, 15 patients had mild illness and 15 patients had moderate/severe illness^22^. As all the patients in the first cohort could not be traced at 12 weeks, in order to carry out the above assays, we recruited a second cohort of patients. Based on the WHO COVID-19 disease classification, 14 patients had mild illness and 6 patients had moderate/severe illness^22^.

In order to compare the antibody responses following infection with one dose of the AZD1222 vaccine, we recruited 20 individuals 4 weeks following vaccination and 73 individuals, 12 weeks following vaccination. We also included serum samples from individuals who had a febrile illness in 2017 and early 2018. Ethical approval was received by the Ethics Review Committee of Faculty of Medical Sciences, University of Sri Jayewardenepura. Informed written consent was obtained from patients.

### Haemagglutination test (HAT) to detect antibodies to the receptor binding domain (RBD)

The HAT was carried out as previously described ^23^. The B.1.1.7 (N501Y), B.1.351 (N501Y, E484K, K417N) and B.1.617.2 versions of the IH4-RBD reagent were produced as described ^23^, but included the relevant amino acid changes introduced by site directed mutagenesis. These variants were titrated in a control HAT with the monoclonal antibody EY-6A (to a conserved class 4 epitope^23,24^) and found to titrate identically with the original version so 100ng (50ul of 2ug/ml stock solution) was used for developing the HAT. The assays were carried out and interpreted as previously described ^25^. The HAT titration was performed using 11 doubling dilutions of serum from 1:20 to 1:20,480, to determine presence of RBD-specific antibodies. The RBD-specific antibody titre for the serum sample was defined by the last well in which the complete absence of “teardrop” formation was observed.

### Surrogate neutralizing antibody test (sVNT) to detect NAbs

The surrogate virus neutralization test (sVNT)^26^, which measures the percentage of inhibition of binding of the RBD of the S protein to recombinant ACE2^26^ (Genscript Biotech, USA) was carried out according the manufacturer’s instructions as previously described by us^9^. Inhibition percentage ≥ 25% in a sample was considered as positive for NAbs.

## Results

### The kinetics of SARS-CoV-2 specific IgG responses in those with natural infection

IgG responses to the S1, S2, RBD and N protein were measured in individuals with COVID-19 at 4 weeks and at 12 weeks since onset of illness and also in serum samples of 15 individuals who had a febrile illness in 2017 and early 2018. At 4 weeks since onset of illness, the highest magnitude of IgG antibody responses was seen for RBD in those with moderate/severe illness, whereas those with mild disease, had the highest responses to S2 (Figure 1A, table 1). Those who had a febrile illness in year 2017 and 2018 (controls), also had detectable antibody levels to S2, but not for other proteins. There was no difference in the antibody levels to S2 in those with mild illness compared to the controls (p=0.13), although those with milder disease had significantly higher antibody levels to S1 and RBD (p<0.0001) and N protein (p=0.0004), than the controls. In those who received a single dose of the AZD1222 vaccine, the IgG responses to the S1 and S2 components of the spike protein were similar, although the levels for the RBD was significantly higher (table 1). As expected, the IgG responses to the N protein was very low, but even lower than for the controls. The antibody levels to S1 (p=0.0002), S2 (p=0.01), RBD (p=0.002) and N (p<0.0001) proteins were significantly different between the three groups of individuals at 4 weeks (Figure 1A).

**Figure 1:**
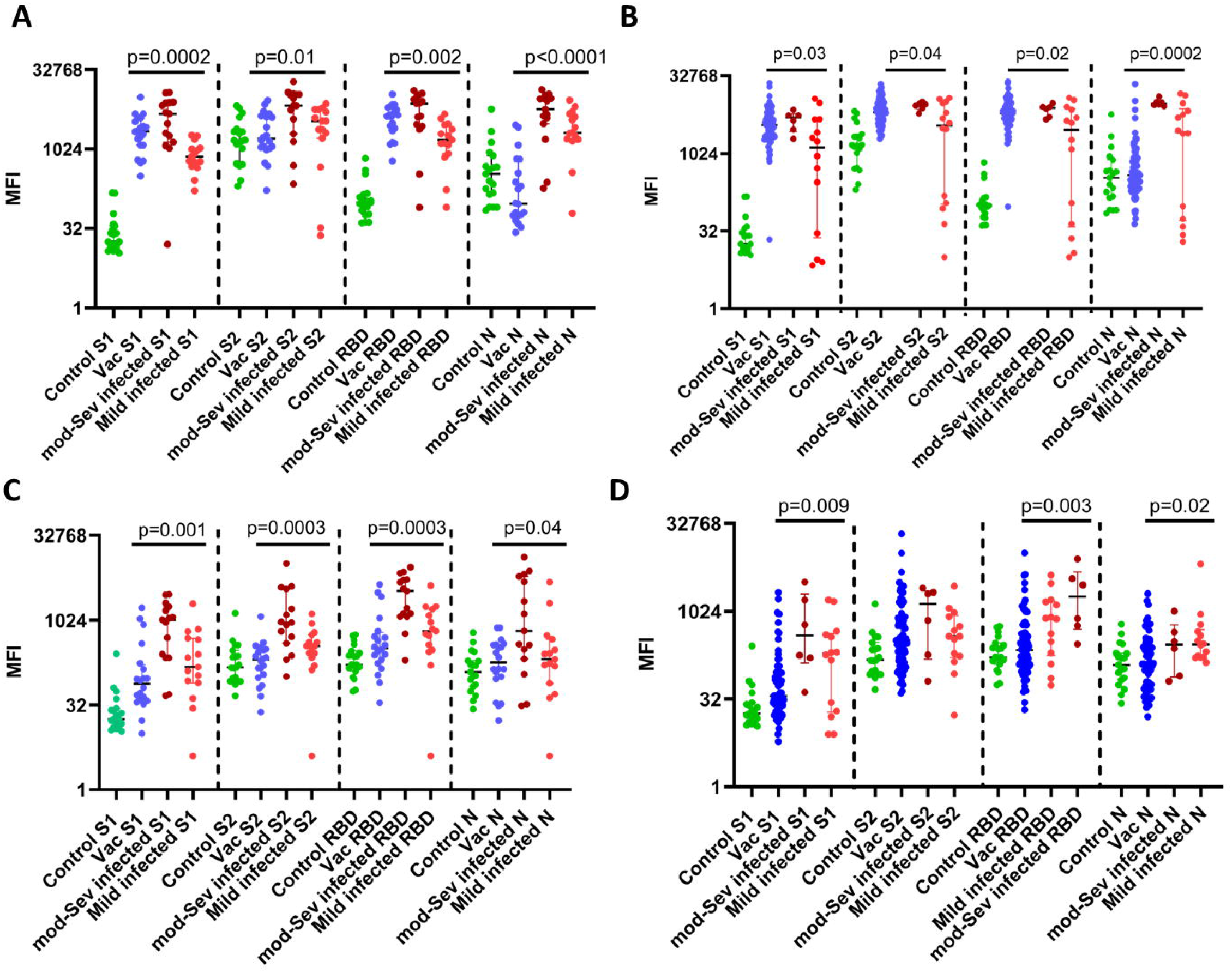
IgG and IgA antibody levels to S1, S2, RBD and N protein of SARS-CoV-2 in individuals following natural infection and following a single dose of the AZD1222 vaccine. Serum IgG antibodies to S1, S2, RBD and N protein were measured by Luminex assays at 4 weeks in those with mild illness (n=15), moderate/severe illness (n=15), vaccinees (n=20) and controls (n=19) (A) and again at 12 weeks in those with mild illness (n=14), moderate/severe illness (n=6), vaccinees (n=73) (B). IgA antibodies were also measured in the above groups at 4 weeks (C) and at 12 weeks (D). The Kruskal-Wallis test was used to determine the difference between the antibody levels between the three different groups (two-tailed). The lines indicate the median and the interquartile range.

**Table 1:**
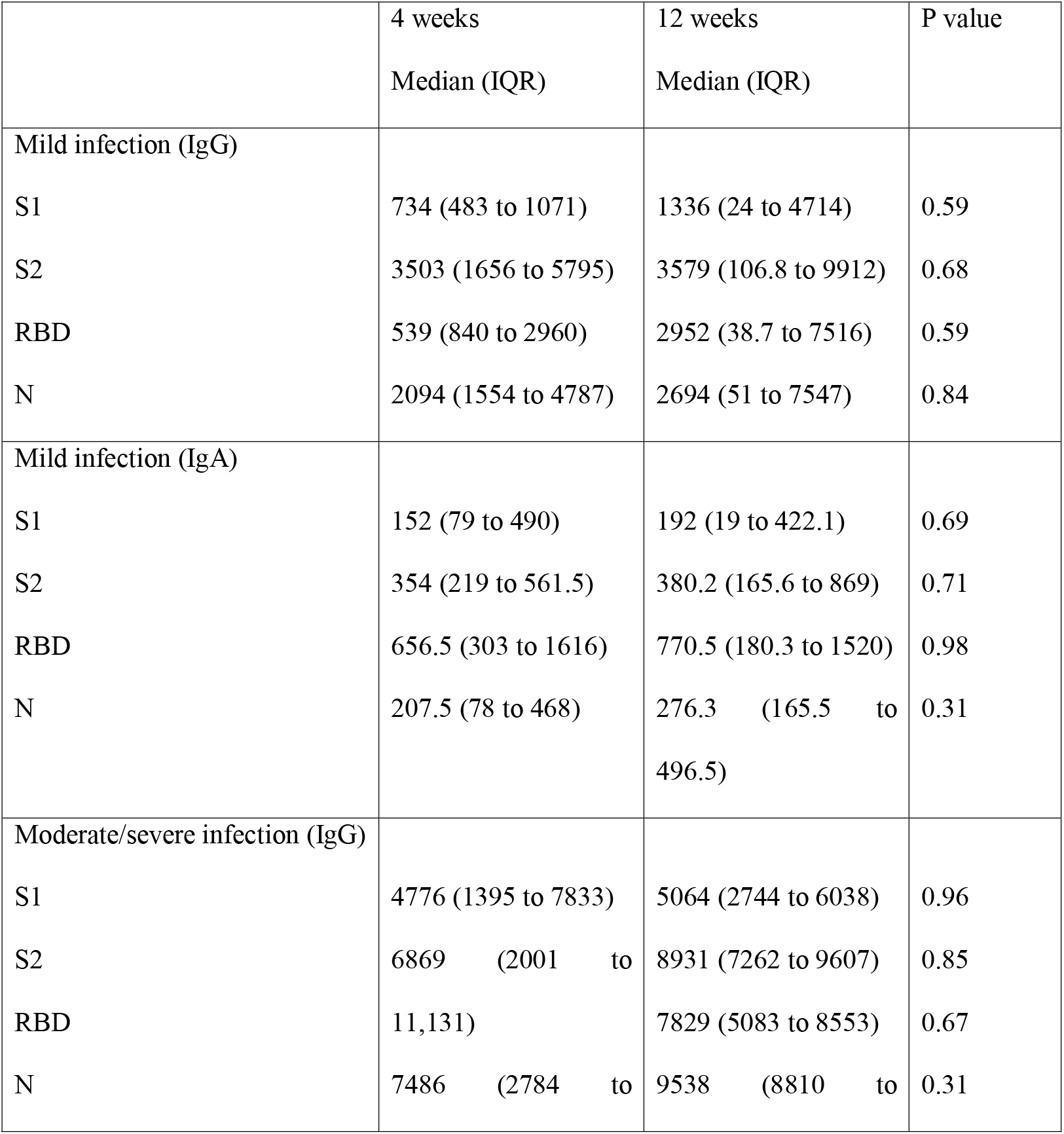
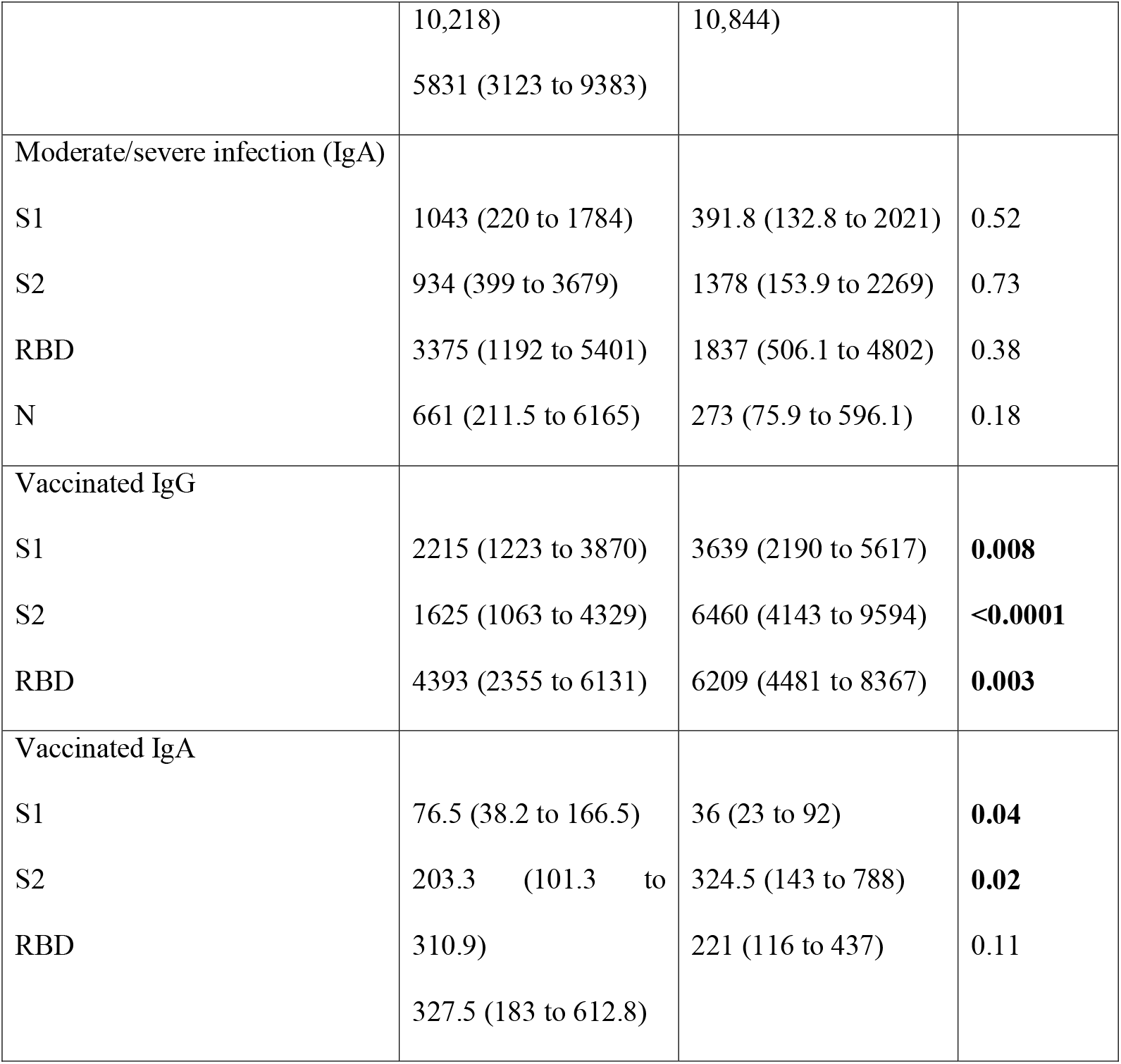
Antibody responses to S1, S2, RBD and N protein of the SARS-CoV-2 in those with varying severity of illness and in those following a single dose of the AZD2221. MFI indicates the median fluorescence intensity.

At 12 weeks since onset of illness, those with moderate/severe illness had the highest responses to N protein, whereas those with mild illness still had the highest responses to S2 (Figure 1B). At both time points for all proteins, those with moderate/severe disease had significantly higher antibody levels than those with milder illness (Table 1). The antibody responses to S1 (p=0.03), S2 (p=0.04), RBD (p=0.02) and N protein (p=0.0002) were significantly different between the those with mild illness, moderate/severe disease and the vaccinees (Figure 1B). From 4 to 12 weeks, the S1 specific antibodies significantly rose in those with mild illness (p=0.004), while there was no significant change in the antibody levels to other proteins at 12 weeks (table 1). Patients who had moderate/severe illness sustained the same levels of antibodies for all four proteins from 4 weeks to 12 weeks. In the vaccinees, from 4 weeks to 12 weeks the IgG levels to S1 (p=0.008), S2 (p<0.0001) and RBD (p=0.003) had significantly increased (table 1).

### The kinetics of SARS-CoV-2 specific IgA responses in those with natural infection

IgA responses to the S1, S2, RBD and N protein were measured in the above individuals with COVID-19 at 4 weeks and at 12 weeks since onset of illness and also in serum samples of 15 individuals who had a febrile illness in 2017 and early 2018. At 4 weeks and 12 weeks of illness, individuals with both mild and moderate/severe illness, had the highest levels of IgA antibodies to the RBD (Figure 1C and 1D). However, those with moderate/severe disease had significantly higher antibody responses to all four proteins when compared to those with mild illness at 4 weeks, but there was no difference at 12 weeks (table 1). Unlike what was observed with SARS-CoV-2 S2 specific IgG responses, those with mild illness had significantly higher IgA responses (p=0.02) but not to N protein (p=0.18) (Figure 2A). As expected, vaccinees had low responses to the N protein, with IgA levels similar to those seen in controls except for IgA to S1, which was higher in the vaccinees (p=0.003). Significant differences of IgA responses were seen in those with mild illness, moderate/severe illness and vaccinees for S1 (p=0.001), S2 (p=0.0003), RBD (p=0.0003) and N protein (p=0.04) (Figure 1C).

**Figure 2:**
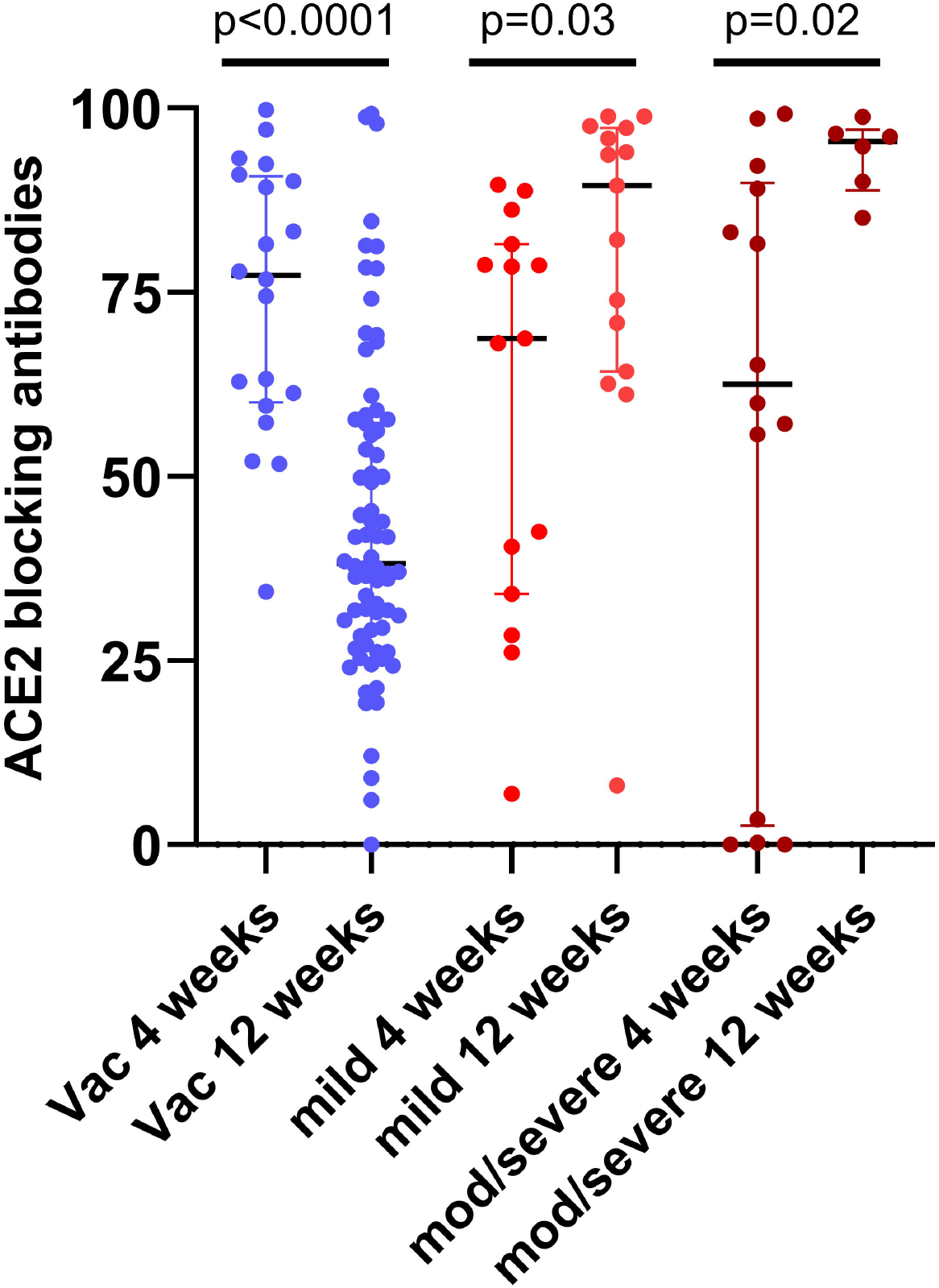
ACE2 receptor blocking antibodies in patients with varying severity of illness and following a single dose of the AZD1222 vaccine. ACE receptor blocking antibodies were measured by the surrogate virus neutralizing test following natural infection at 4 weeks in those with mild illness (n=14) and moderate/severe illness (n=15) and at 12 weeks in those with mild (n=14) and moderate/severe illness (n=6). Antibodies were also measured at 4 weeks (n=20) and 12 weeks (n=73) in vaccinees following a single dose of AZD2221. Mann-Whitney test (two tailed) was used to determine the differences between antibody levels as 4 weeks and 12 weeks. The lines indicate the median and the interquartile range.

There was no difference in IgA levels to any of the proteins at 4 weeks compared to 12 weeks in patients with mild illness or with moderate/severe illness (table 1). However, significant differences were seen between the three groups to S1 (p=0.009), RBD (p=0.003) and N protein (p=0.02), but not for S2 (p=0.55) (Figure 1D).

### ACE2 receptor blocking antibodies following natural infection and one dose of AZD1222

Due the lack of BSL-3 facilities to measure neutralizing antibodies, we used a surrogate test to measure the inhibition of binding of antibodies in patient sera to the ACE2 receptor. This was shown to be 100% specific in the Sri Lankan population, with none of the sera of individuals collected in 2017 and 2018, giving a positive response ^9^. The ACE2 blocking antibodies were significantly higher in those with moderate to severe illness, when compared to those with mild illness at 4 weeks (p=0.03) and at 12 weeks (p=0.03) as reported previously^9^ (Figure 2). In addition, the ACE2 receptor blocking antibodies significantly increased from 4 weeks to 12 weeks in those with moderate/severe illness (p=0.02) and in those with mild illness (p=0.03) (Figure 2). However, in those who received a single dose of the vaccine, the ACE2 blocking antibodies significantly reduced (p<0.0001) from levels at 4 weeks (median 77.32, IQR 60.05 to 90.77 % of inhibition) to 12 weeks (median 38.17, IQR 28.95 to 57.28 % of inhibition).

### Antibodies to the Receptor Binding Domain of the spike protein, including variants, measured by the Haemagglutination test (HAT)

We previously evaluated the usefulness of the HAT assay in determining antibody responses to the RBD of the SARS-CoV-2, wild type (WT) virus, B.1.1.7 variant and the B.1.351 variants at 4 weeks following a single dose of the AZD1222 vaccine and had also evaluated this assay in naturally infected individuals in Sri Lanka ^27^. In this study, we proceeded to investigate the differences in the antibody responses to the RBD in those with natural infection at 4- and 12- weeks following infection, and after a single dose of the AZD1222 vaccine. The antibody responses to the WT, B.1.1.7, B.1.351 and B.1.617.2 were measured.

In those with mild illness, at 4 weeks from onset of illness the median antibody titre to the WT was 160 (IQR 80 to 320), B.1.1.7 was 120 (IQR 70 to 320), B.1.351 was 10 (IQR, 0 to 80) and for B.1.617.2 it was 40 (IQR 20 to 80). At 12 weeks following the onset of illness, although there was a slight reduction in the antibody titres to the WT (p=0.91) and B.1.617.2 (0.61), this was not statistically significant (Figure 3A). In those with moderate/severe illness at 4 weeks from onset of illness the median antibody titre to the WT was 1280 (IQR 160 to 1280), B.1.1.7 was 640 (IQR 160 to 1280), B.1.351 was 40 (IQR, 0 to 160) and for B.1.617.2 it was 320 (IQR 80 to 1280) (Figure 3B). There was no significant difference between the antibody titres for the WT compared to B.1.1.7 (p=0.12), but clearly differed for B.1.315 (p<0.0001) and B.1.617.2 (p=0.004). Although the antibody titres for the WT and all the variants reduced from 4 to 12 weeks in those with moderate/severe illness, this was not statistically significant (Figure 3B).

**Figure 3:**
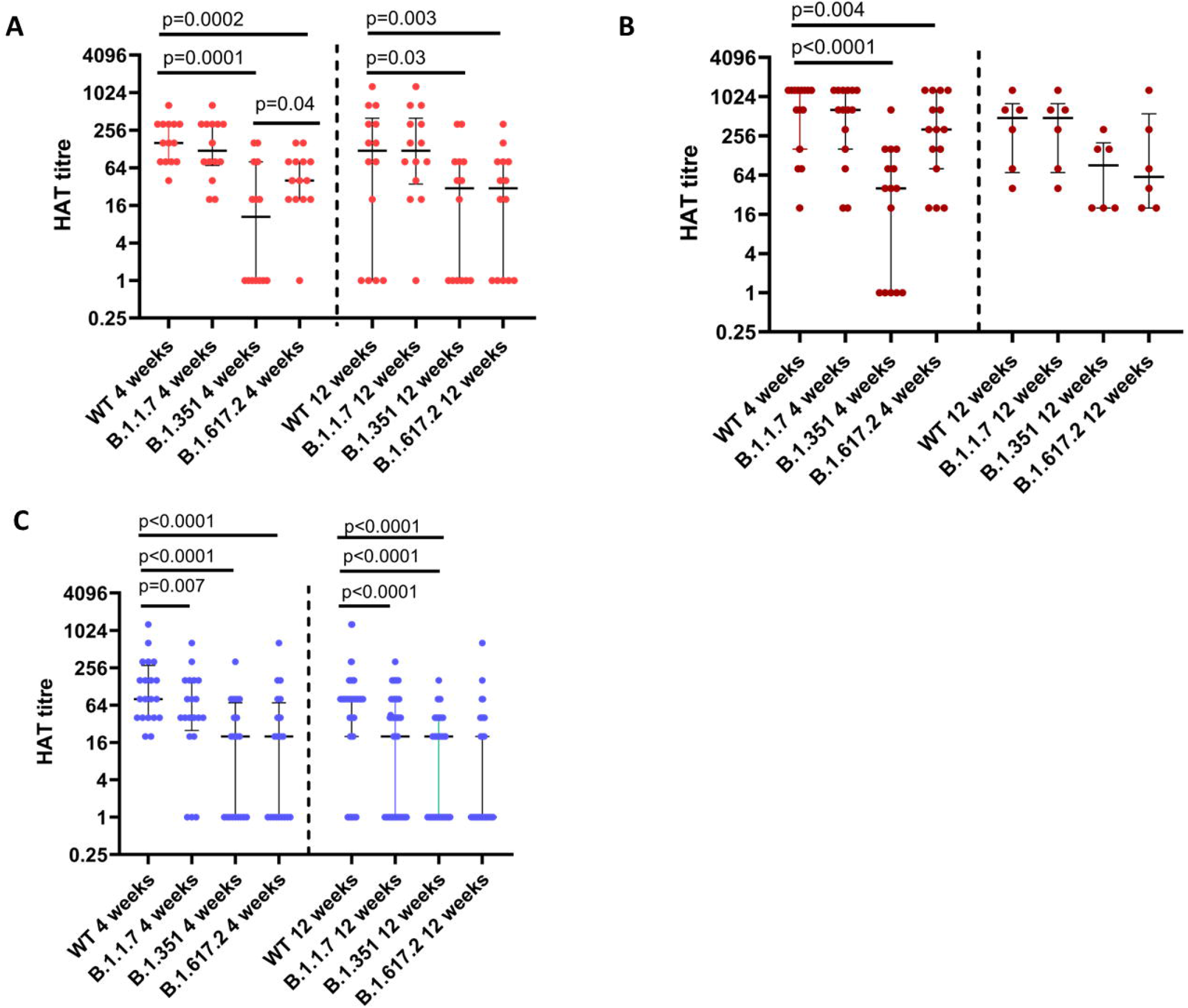
Comparison of antibody titres to RBD of the SARS-CoV-2 using the HAT assay in those with varying severity of infection and in vaccinees. Antibody titres were measured individuals with mild illness (n=14) to the WT, B.1.1.7, B.1.351 and B.1.617.2 at 4 weeks and 12 weeks since onset of illness (A), in those with moderate/severe illness at 4 weeks (n=15) and 12 weeks (n=6) since onset of illness (B) and in those who received one dose of AZD1222 vaccine at 4 weeks (n=16) and 12 weeks (n=73) following the vaccine (C). The difference between antibody titres to WT, B.1.1.7, B.1.351 and B.1.617.2 was determined using the Wilcoxon paired t test (two tailed) and the differences between antibody titres at 4 weeks and 12 weeks was determined using the Mann-Whitney test (two tailed). The lines indicate the median and the interquartile range.

At 4 weeks following a single dose of the vaccine, the median antibody titre to the WT was 80 (IQR 40 to 280), B.1.1.7 was 40 (IQR 20 to 160), B.1.351 was 20 (IQR, 0 to 70) and for B.1.617.2 it was 20 (IQR 0 to 70) (Figure 3C). At 12 weeks following a single dose of the vaccine, the antibody titre for WT was 80 (IQR 20 to 80), for B.1.1.7 it was 20 (IQR 0 to 80), for B.1.351 it was 20 (0 to 40) and for B.1.617.2 it was 0 (IQR 0 to 20). From 4 to 12 weeks, although there was no significance difference of the antibody titres of the RBD of the WT (p=0.05), B.1.351 (p=0.54) and B.617.2 (p=0.07), the antibody titres to B.1.1.7 significantly reduced (p=0.02) (Figure 3B). As previously described by us at 4 weeks following vaccination, the HAT titres were significantly lower to the B.1.1.7 (p=0.007), B.1.351 (<0.0001) and for B.1.617.2 (p<0.0001). However, there was no significance difference in antibody titres between B.1.351 and B.1.617.2 (p=0.43). At 12 weeks again the HAT titres were significantly lower to the B.1.1.7 (p<0.0001), B.1.351 (<0.0001) and for B.1.617.2 (p<0.0001) and no difference between antibody titres to B.1.351 and B.1.617.2.

Antibodies to the RBD were significantly different between those with mild illness, moderate/severe illness and with those with a single dose of the vaccine at 4 weeks (p=0.004) and at 12 weeks (p=0.02) (Figure 4A). This difference was also seen for the B.1.1.7 at 4 weeks between those with mild illness, moderate/severe illness and with those with a single dose of the vaccine at 4 weeks (p=0.0006) and at 12 weeks (p<0.0001) (Figure 4B) and for B.1.617.2 at 4 weeks (p=0.0002) and at 12 weeks (p=0.0004) (Figure 4C). However, there was no difference between the antibody titres to the B.1.351 between those with mild, moderate/severe illness and vaccinees at 4 weeks (p=0.36), but a significant difference was seen at 12 weeks (p=0.02) (Figure 4D).

**Figure 4:**
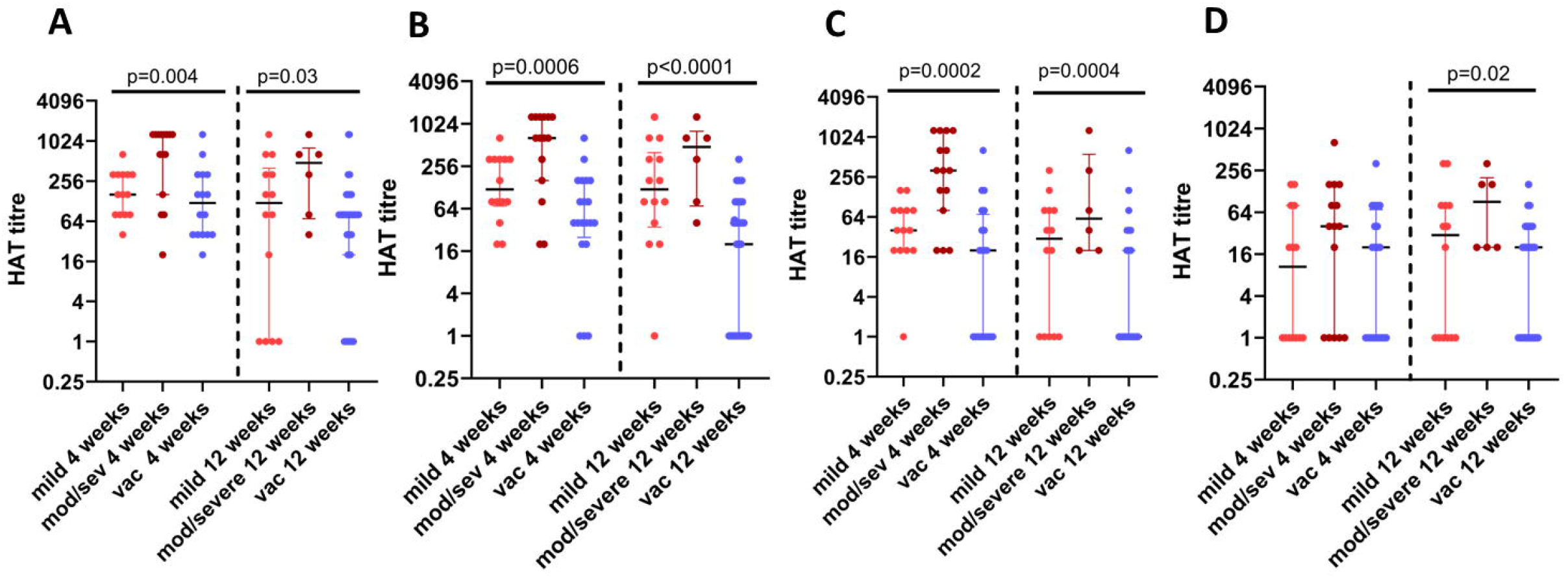
Comparison of antibody titres to the RBD of the SARS-CoV-2 using the HAT assay for the wild type and for variants. Antibody titres were measured in patients with mild illness (n=14), moderate/severe illness (n=15) from 4 weeks since onset of illness and in those who received one dose of AZD1222 vaccine at 4 weeks (n=16), and again at 12 weeks in those who developed mild illness (n=14), moderate/severe illness (n=6) and in those who received 1 dose of AZD1222 vaccine (n=73), for the WT (A), B.1.1.7 (B), B.1.617.2 (C) and B.1.351 (D). The Kruskal-Wallis test was used to determine the difference between the antibody levels between the three different groups (two-tailed). The lines indicate the median and the interquartile range.

## Discussion

In this study we have investigated the kinetics of IgG and IgA responses to S1, S2, RBD and N protein, ACE2 receptor blocking antibodies and antibodies against SARS-CoV-2 variants, in individuals at 4 and 12 weeks following natural infection and in those who had a single dose of the AZD2221. Based on the Luminex assays, IgG and IgA levels to S1, S2, RBD and N, had increased from 4 weeks to 12 weeks in those with mild illness and in the vaccinees, although the increase was only significant in the vaccinees. In the vaccinees, the most significant rise was seen for the S2 subunit, while in those with mild illness the rise was seen for IgG antibodies for the RBD. In those with moderate/severe illness, while there was no change in the IgG responses from 4 to 12 weeks, responses to the N protein had increased although this was not significant. Therefore, the kinetics of antibody responses to S1, S2, RBD and N appears to vary based on the severity of natural infection and also appeared to be different in vaccinees. Interestingly, blood samples of those who had a febrile illness in 2017 and 2018 also showed IgG and IgA responses to the S2 subunit, suggesting the presence of S2 subunit cross-reactive antibodies, in these donors as previously seen in other studies^13,14^. Following a single dose of the AZD2221 vaccine, the antibodies against S2 appears to continue to rise from 4 to 12 weeks, possibly due to stimulation of pre-existing cross-reactive memory B cell responses to the S2 subunit^14^.

SARS-CoV-2 specific IgA antibodies have shown to be generated during early illness and have potent neutralizing ability^15^. IgA antibodies to the RBD have shown to develop earlier than IgG and while some studies have shown that serum IgA does not associate with clinical disease severity^15^, in other studies, patients who developed severe disease were shown to have higher levels of virus specific IgA^28^. Serum IgA was shown to activate neutrophils, thereby leading to production of increased levels of inflammatory mediators possibly leading to disease pathogenesis^16^. We found that at 4 weeks of illness, those with moderate/severe illness had significantly higher serum IgA to S1, S2, RBD and N compared to those with mild illness, but these high levels of IgA declined and there was no differences between these two groups at 12 weeks since onset of illness. Vaccinees had several fold lower IgA antibodies to all the SARS-CoV-2 proteins tested than those with mild and moderate/severe illness at 4 weeks and 12 weeks. The importance of serum IgA in preventing re-infection is currently unknown and if those with lower IgA have reduced protection is currently unknown.

Although the IgG antibodies to S1, S2 and the RBD rose from 4 to 12 weeks in the vaccinees, the ACE2 receptor blocking antibodies, which were shown to correlate with neutralizing antibodies significantly decreased^26^. The HAT assay, which also measures antibodies to the RBD and has shown to correlate well with the ACE2 receptor blocking assay and with neutralizing antibodies^23,27^, also showed that the RBD binding antibodies decreased from 4 to 12 weeks in the vaccinees. This suggests that although antibodies to RBD, S1 and S2 have increased in vaccinees from 4 to 12 weeks, they might not be neutralizing antibodies, possibly through targeting other epitopes in these regions.

Apart from assessing antibodies to the RBD to the wild type, we assessed the antibodies to three other VOCs, B.1.1.7, B.1.351 and B.1.617.2. At 4 weeks following vaccination, the vaccinees had similar levels of antibodies to the RBD of WT as those with mild illness, the levels were significantly less for B.1.1.7 and for B.1.617.2. The antibody levels among vaccinees were similar to B.1.351 and B.1.617.2, showing equal reduction compared to antibody binding to the RBD of the WT. Following vaccination, these levels further declined at 12 weeks to VOCs but not to the WT, showing that a single dose of the AZD2221 was likely to offer less protection against VOCs. In fact, it has been shown that one dose of AZD2221 is only 33% effective in preventing symptomatic disease with B.1.617.2, 3 weeks following the first dose^29^. The efficacy of a single dose against B.1.617.2 is likely to decline further by 12 weeks, as the antibodies to RBD further waned. However, the efficacy of two doses of AZD2221 against hospitalization has been shown to be 92%, while for Pfizer-BioNTech was 96%^30^. Therefore, in countries which have outbreaks due to VOCs, especially B.1.617.2, it would be prudent to encourage second doses to increase efficacy as recommended. Interestingly, although those with mild or moderate/severe illness also had a marked reduction in antibodies to the RBD of B.1.351, they had significantly higher levels of antibodies to the RBD of B.617.2 at 4 weeks compared to B.1.351. However, by 12 weeks the antibody levels to both B.1.351 and B.1.617.2 were similar. Therefore, B.1.617.2 had less immune evasion than B.1.351 in those who were naturally infected, at least during early convalescence.

In summary, we have investigated the kinetics and differences in IgG and IgA antibody responses to the S1, S2, RBD and N in those with varying severity of infection and vaccinees who received a single dose of AZD2221, which showed that vaccinees had significantly less IgA to SARS-CoV-2, but comparable IgG responses those with natural infection. However, following a single dose, vaccinees had reduced antibody levels to the VOCs, which further declined with time.

## Data Availability

All data is available within the manuscript and the figures.

## Acknowledgement

We are grateful to the World Health Organization, UK Medical Research Council and the Foreign and Commonwealth Office for support. T.K.T. is funded by the Townsend-Jeantet Charitable Trust (charity number 1011770) and the EPA Cephalosporin Early Career Researcher Fund. A.T. are funded by the Chinese Academy of Medical Sciences (CAMS) Innovation Fund for Medical Science (CIFMS), China (grant no. 2018-I2M-2-002).

## Notes

### Competing Interest Statement

The authors have declared no competing interest.

### Author Declarations

Ethical approval was received by the Ethics Review Committee of Faculty of Medical Sciences, University of Sri Jayewardenepura, Sri Lanka. Informed written consent was obtained from patients.

## References

1 Medicine, J. H. U. a. Coronavrus Resource Centre, <https://coronavirus.jhu.edu/> (2021).

2 Davies, N. G. et al. Increased mortality in community-tested cases of SARS-CoV-2 lineage B.1.1.7. Nature 593, 270–274, doi:10.1038/s41586-021-03426-1 (2021).

3 England, P. H. SARS-CoV-2 variants of concern and variants under investigation in England. Report No. 15, 77 (Public Health England, 2021).

4 Ritchie H. O.-O. E., Beltekian D., Mathieu E., Hasell J., Macdonald B., Giattino C., Appel C., Lucas Rodés-Guirao, Rose M. (OurWorldInData.org, 2021).

5 Hansen, C. H., Michlmayr, D., Gubbels, S. M., Molbak, K. & Ethelberg, S. Assessment of protection against reinfection with SARS-CoV-2 among 4 million PCR-tested individuals in Denmark in 2020: a population-level observational study. Lancet 397, 1204–1212, doi:10.1016/S0140-6736(21)00575-4 (2021).

6 Sabino, E. C. et al. Resurgence of COVID-19 in Manaus, Brazil, despite high seroprevalence. Lancet 397, 452–455, doi:10.1016/S0140-6736(21)00183-5 (2021).

7 Zhou, D. et al. Evidence of escape of SARS-CoV-2 variant B.1.351 from natural and vaccine-induced sera. Cell 184, 2348–2361 e2346, doi:10.1016/j.cell.2021.02.037 (2021).

8 Chen, X. et al. Disease severity dictates SARS-CoV-2-specific neutralizing antibody responses in COVID-19. Signal Transduct Target Ther 5, 180, doi:10.1038/s41392-020-00301-9 (2020).

9 Jeewandara, C. et al. SARS-CoV-2 neutralizing antibodies in patients with varying severity of acute COVID-19 illness. Sci Rep 11, 2062, doi:10.1038/s41598-021-81629-2 (2021).

10 Brochot, E. et al. Anti-spike, Anti-nucleocapsid and Neutralizing Antibodies in SARS-CoV-2 Inpatients and Asymptomatic Individuals. Front Microbiol 11, 584251, doi:10.3389/fmicb.2020.584251 (2020).

11 Demers-Mathieu, V. et al. Difference in levels of SARS-CoV-2 S1 and S2 subunits-and nucleocapsid protein-reactive SIgM/IgM, IgG and SIgA/IgA antibodies in human milk. J Perinatol 41, 850–859, doi:10.1038/s41372-020-00805-w (2021).

12 Nguyen-Contant, P. et al. S Protein-Reactive IgG and Memory B Cell Production after Human SARS-CoV-2 Infection Includes Broad Reactivity to the S2 Subunit. mBio 11, doi:10.1128/mBio.01991-20 (2020).

13 Ng, K. W. et al. Preexisting and de novo humoral immunity to SARS-CoV-2 in humans. Science 370, 1339–1343, doi:10.1126/science.abe1107 (2020).

14 Fraley, E. et al. Cross-reactive antibody immunity against SARS-CoV-2 in children and adults. Cellular & molecular immunology, doi:10.1038/s41423-021-00700-0 (2021).

15 Sterlin, D. et al. IgA dominates the early neutralizing antibody response to SARS-CoV-2. Science translational medicine 13, doi:10.1126/scitranslmed.abd2223 (2021).

16 Bartsch, Y. C. et al. Humoral signatures of protective and pathological SARS-CoV-2 infection in children. Nature medicine 27, 454–462, doi:10.1038/s41591-021-01263-3 (2021).

17 Abu-Raddad, L. J., Chemaitelly, H., Butt, A. A. & National Study Group for, C.-V. Effectiveness of the BNT162b2 Covid-19 Vaccine against the B.1.1.7 and B.1.351 Variants. The New England journal of medicine, doi:10.1056/NEJMc2104974 (2021).

18 Hall, V. J. et al. COVID-19 vaccine coverage in health-care workers in England and effectiveness of BNT162b2 mRNA vaccine against infection (SIREN): a prospective, multicentre, cohort study. Lancet 397, 1725–1735, doi:10.1016/S0140-6736(21)00790-X (2021).

19 Rubin, R. COVID-19 Vaccines vs Variants-Determining How Much Immunity Is Enough. JAMA : the journal of the American Medical Association 325, 1241–1243, doi:10.1001/jama.2021.3370 (2021).

20 Tauh. T M. M., Meyler, P., Lee, S.M. An updated look at the 16-week window between doses of vaccines in BC for COVID-19. BC Medical Journal 63, 102–103 (2021).

21 Campillo-Luna, J., Wisnewski, A. V. & Redlich, C. A. Human IgG and IgA responses to COVID-19 mRNA vaccines. medRxiv, 2021.2003.2023.21254060, doi:10.1101/2021.03.23.21254060 (2021).

22 WHO. Clinical management of severe acute respiratory infection when novel coronavirus (2019-nCoV) infection is suspected: interim guidance. (WHO, 2020).

23 Townsend, A. et al. A haemagglutination test for rapid detection of antibodies to SARS-CoV-2. Nature Communications, 2020.2010.2002.20205831, doi:10.1101/2020.10.02.20205831 (2020).

24 Zhou, P. et al. A pneumonia outbreak associated with a new coronavirus of probable bat origin. Nature 579, 270–273, doi:10.1038/s41586-020-2012-7 (2020).

25 Jeewandara, C. et al. Antibody and T cell responses to a single dose of the AZD1222/Covishield vaccine in previously SARS-CoV-2 infected and naïve health care workers in Sri Lanka. medRxiv, 2021.2004.2009.21255194, doi:10.1101/2021.04.09.21255194 (2021).

26 Tan, C. W. et al. A SARS-CoV-2 surrogate virus neutralization test based on antibody-mediated blockage of ACE2-spike protein-protein interaction. Nature biotechnology 38, 1073–1078, doi:10.1038/s41587-020-0631-z (2020).

27 Kamaladasa, A. et al. Comparison of two assays to detect IgG antibodies to the receptor binding domain of the SARSCoV2 as a surrogate marker for assessing neutralizing antibodies in COVID-19 patients. Int J Infect Dis, doi:10.1016/j.ijid.2021.06.031 (2021).

28 Yu, H. Q. et al. Distinct features of SARS-CoV-2-specific IgA response in COVID-19 patients. The European respiratory journal 56, doi:10.1183/13993003.01526-2020 (2020).

29 Iacobucci, G. Covid-19: Single vaccine dose is 33% effective against variant from India, data show. BMJ (Clinical research ed 373, 1346, doi:10.1136/bmj.n1346 (2021).

30 England, P. H. (Public Health England, 2021).

